# Comparison of Glycemic Control between Intensive Insulin Regimen and Continuous Subcutaneous Insulin infusion: A Meta-Analysis Report of Type-1 Diabetics from Randomized Controlled Trials

**DOI:** 10.1101/2020.06.01.20119693

**Authors:** Kamran Mahmood Ahmed Aziz, Abdullah Othman

## Abstract

Achieving glycemic control and targets are challenging in type-1 diabetes management. To achieve this, intensive insulin therapy or multiple daily injections (MDI) and continuous subcutaneous insulin infusion (CSII) or pump therapy have been used in various health care settings. However, there has been a debate on their superiority. Some of researchers have recommended MDI, while others SCII. We compared MDI with CSII by a literature search. We have conducted mata-analysis for MDI and CSII on ten randomized controlled trials on 809 type-1 diabetics 809, MDI (N = 394) or CSII (N = 415). Heterogeneity between trials was quantified by *conventional Q-statistic* (Cochran’s heterogeneity statistic) and Higgins *I^2^* statistic with 0-40% representing negligible heterogeneity, 30-60% moderate heterogeneity, 50-90% substantial heterogeneity and 75-100% considerable heterogeneity. tau-squared (τ^**2**^) was used to observe between-study random-effects variance. Meta Analyst software was used to analyze the data and to conduct meta-analysis. SPSS was used to analyze HbA1c student’s t-test for MDI and CSII. A random-effect analysis ((DerSimonian-Laird method) performed on ten studies found that the percentage of HbA1c was lower in patients receiving CSII compared with those receiving MDI; standardized mean difference (SMD) was 0.441, 95% confidence interval 0.267 to 0.616, p < 0.001; equivalent to a difference of 0.39%, favoring CSII. *I^2^* statistic was 20.9; τ^**2**^ = 0.016; Q = 11.378 with df = 9, indicating that heterogeneity was not significant (heterogeneity p-value = 0.251). Patients on CSII demonstrated significantly lower values (8.2±0.72 versus 7.73±0.72; p-value < 0.001 respectively). This statistical and meta-analysis favors the usage of insulin pump therapy. We concluded that patient centered approach should be used while selecting the patients for insulin pump (CSII) or MDI.

## Introduction

There has been a debate that continuous subcutaneous insulin infusion (CSII) or pump therapy (first introduced in 1970s) is superior to insulin injections including multiple daily injections (MDI), intensive insulin therapy or the basal bolus therapy. Furthermore, some studies have demonstrated that insulin dosages were less in CSII with better patient satisfaction [1 2]. However, there are several other studies with some conflicting results and some authors have concluded that both are equally effective in term of reduction of glycated hemoglobin (HbA1c) [3–5]. The main outcome of these studies was HbA1c, which shows the control of diabetes for the past two months. HbA1c is important, as if this worsens, diabetes complications initiates and progress [6]. Under this debate, we collected randomized clinical trials (RCTs) conducted on type-1 diabetic patients comparing the HbA1c results between MDI and CSII and conducted meta-analysis.

## Materials and methods

PRISMA guidelines were used for reporting of individual patient data meta-analyses [7]. We performed internet database survey (PubMed, Google Scholar) and reviewed literature. Only randomized controlled trials (RCTs) on type-1 diabetic patient were included. Observational studies, reviews, surveys, and short term studies (less than two months) were excluded. Also studies with incomplete data and those studies which did not provide complete data details (such as mean ±SD or the numbers randomized/exact number of subjects) were excluded from meta-analysis. HbA1c mean ± SD was calculated for MDI and CSII. Heterogeneity between trials was quantified by *conventional Q-statistic* (Cochran’s heterogeneity statistic) and Higgins *I^2^* statistic (the degree of inconsistency in the results between studies or the percentage of variability in effect due to heterogeneity rather than sample error) with 0–40% representing negligible heterogeneity, 30–60% moderate heterogeneity, 50–90% substantial heterogeneity and 75–100% considerable heterogeneity. Additionally, tau-squared (τ^**2**^), estimates for the between-study random-effects variance was calculated as well. Standardized statistical techniques and Meta Analyst software was used to analyze the data and to conduct meta-analysis [8 – 12]. Data was also entered in SPSS to find mean HbA1c differences (t-test) for MDI and CSII. A random-effect analysis was performed on these studies to find out overall effect measure.

## Results

According to inclusion criteria, ten studies were identified as RCT on type-1 diabetics, with 809 patients randomized to receive either MDI (N = 394) or CSII (N = 415). Table-1 demonstrates details and characteristics of the trials included in the meta-analysis [13-22]

**Table-1.** Randomized controlled trail name, year, duration, number of participants with HbA1c mean ± SD for MDI and CSII.

| Name of Study<br>(Randomized trial) | year | MDI<br>(N) | MDI<br>HbA1c<br>Mean ±SD | CSII<br>(N) | CSII (N)<br>HbA1c<br>Mean ±SD | Study<br>Duration |
| --- | --- | --- | --- | --- | --- | --- |
| Hirsch IB, et al. | 2005 | 50 | 7.3 ± 0.7 | 50 | 7.1 ± 0.8 | 10 weeks |
| Bolli GB, et al. | 2009 | 30 | 7.8 ± 0.6 | 28 | 7.7 ± 0.7 | 24 weeks |
| Hanaire-Broutin, HE et al. | 2000 | 9 | 8.24 ± 0.77 | 32 | 7.89 ± 0.77 | 16 weeks |
| Alemzadeh R, et al. | 2004 | 40 | 8.2 ± 0.9 | 40 | 7.8 ± 0.8 | 12 months |
| Skogsberg L, et al. | 2008 | 38 | 6.7 ± 0.5 | 34 | 6.5 ± 0.4 | 24 months |
| Weintrob N, et al. | 2004 | 12 | 8.2 ± 0.8 | 11 | 8 ± 0.8 | 14 weeks |
| Reznik Y, et al. | 2014 | 163 | 8.6 ± 1.1 | 168 | 7 ± 1.2 | 24 weeks |
| Doyle EA, et al. | 2004 | 16 | 8.1 ± 1.2 | 16 | 7.2 ± 1 | 16 weeks |
| Lepore G, et al. | 2004 | 24 | 9 ± 1.3 | 24 | 8 ± 1 | 12 months |
| Marshall SM, et al. | 1987 | 12 | 9 ± 0.4 | 12 | 9.2 ± 0.5 | 24 weeks |

Figure-1 shows a forest plot and results of aggregate meta-analysis with the effect size of all ten studies, their confidence intervals (95% CI), and the summary with overall effect measure for the mean HbA1c difference between MDI and CSII.

**Fig-1.**
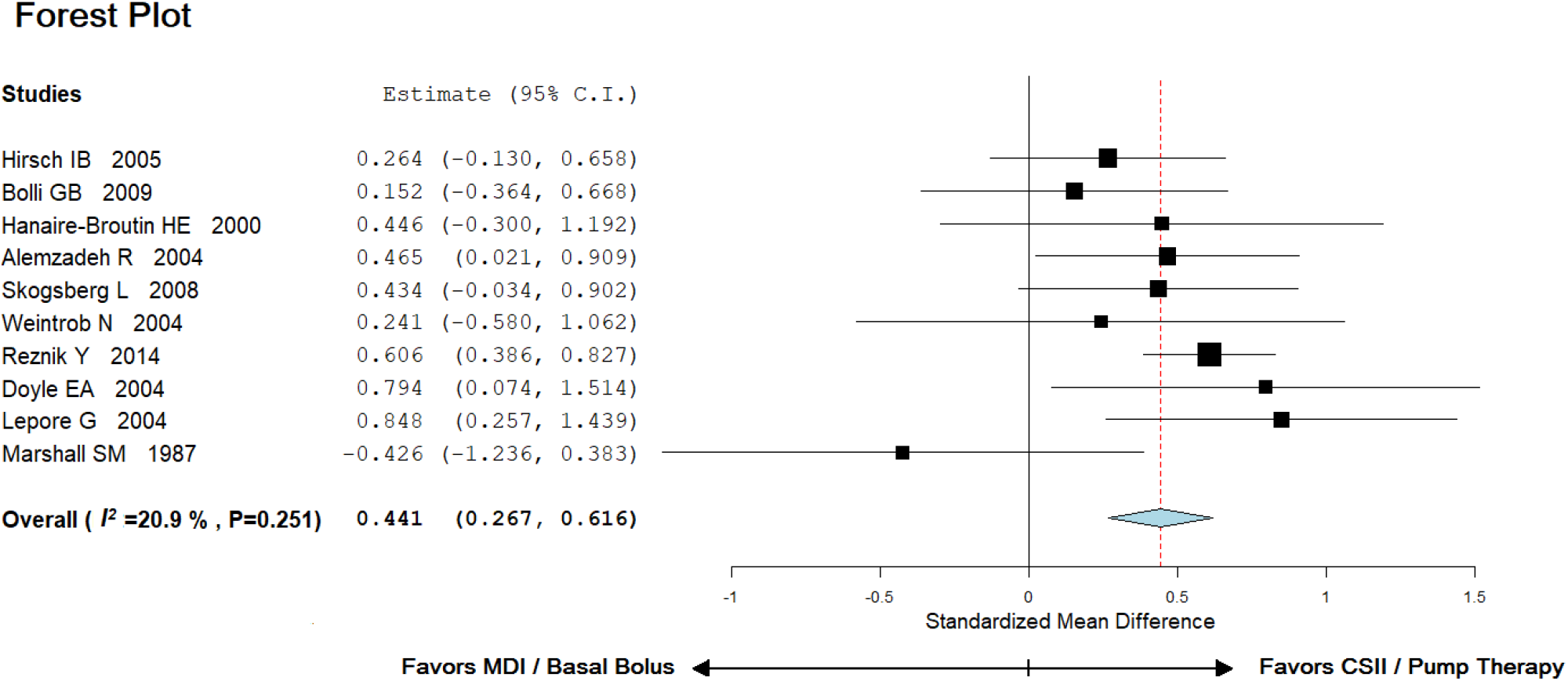
Forest Plot results of random effect meta-analysis model (DerSimonian-Laird random effects method) with standardized mean differences (SDM), 95% confidence intervals for percentage of glycated hemoglobin (HbA1c%) compared with insulin pump (CSII) versus MDI or basal bolus therapy(*SMD = 0.441 (95% CI 0.267 to 0.616) I^2^ = 20.9; τ^**2**^ = 0.016; Q = 11.378 df = 9; p = 0.251)*

A random-effect analysis ((DerSimonian-Laird method) performed on ten studies found that the percentage of glycated Haemoglobin (HbA1c) was lower in patients receiving continuous subcutaneous insulin infusion compared with those receiving insulin injections; standardized mean difference (SMD) was 0.441, 95% confidence interval 0.267 to 0.616, p < 0.001; equivalent to a difference of 0.39%, favoring CSII. *I^2^* statistic was 20.9; τ^**2**^ = 0.016; Q = 11.378 with df = 9, indicating that heterogeneity was not significant (heterogeneity p-value = 0.251). When mean HbA1c values of MDI and CSII were compared, patients on CSII demonstrated significantly lower values (8.2±0.72 versus 7.73±0.72; p-value < 0.001 respectively). This statistical and meta-analysis favors the usage of insulin pump therapy.

## Discussion conclusion and recommendations

Successful management of diabetes requires diabetes self-management education (DSME) and a team work. Selecting the patient for specific management strategies is an art. While selecting the patient for MDI or CSII, several factors should be considered, such as patient’s age, his glycemic profile, HbA1c, dietary pattern, other comorbid complications and a history of hypoglycemia (such as late night, early morning or daytime hypoglycemia). Patient counselling is also an essential aspect of diabetes management prior to starting or selecting specific insulin therapies or regimens, such as MDI or CSII. Although different studies in medical literature have given different conclusions, however, our meta-analysis favors the use of insulin pump in type-1 diabetics for better glycemic control. Some studies conducted in past have also concluded that insulin pump provides only satisfaction to the patients and that glycemic control was equally effective with MDI or CSII [17]. While on the other hand, some studies have reported lower risk of hypoglycemia with CSII [13]. Conversely, other authors have proved that the incidence of hypoglycemia was similar with CSII and MDI [14, 22]. Under this discussion and meta-analysis, physicians and diabtologists should use patient centered approach for managing hyperglycemia in type-1 diabetics [23, 24]. Patient’s selection for optimal therapies remains the top priority which can be achieved by DSME and counselling. Furthermore, cost effectiveness should also be considered while selecting MDI and CSII. Further studies at multicenter level are required to confirm the findings of the current study.

## Data Availability

Data fro RCT

## References

1. Pickup JC, Keen H, Parsons JA, Alberti KGMM. Continuous subcutaneous insulin infusion: an approach to achieving normoglycae-mia. BMJ 1978;i:204–7.

2. Nathan DM, Lou P, Avruch J. Intensive conventional and insulin pump therapy in adult type 1 diabetes. A crossover study. Ann Intern Med 1982;97:31–6.

3. Jeitler K, Horvath K, Berghold A, Gratzer TW, Neeser K, Pieber TR, Siebenhofer A. Continuous subcutaneous insulin infusion versus multiple daily insulin injections in patients with diabetes mellitus: systematic review and meta-analysis.

4. American Diabetes Association. Continuous subcutaneous insulin infusion. Diabetes Care 2004; 27: 110.

5. NICE. Guidance on the use of continuous subcutaneous insulin infusion for diabetes. Technology appraisal guidance no. 57. London: National Institute for Clinical Excellence, 2003.

6. Diabetes Control and Complications Trial Research Group. The effect of intensive treatment of diabetes on the development and progression of long-term complications in insulin-dependent diabetes mellitus. N Engl J Med 1993;329:977–86.

7. Moher D, Liberati A, Tetzlaff J, Altman DG. Preferred reporting items for systematic reviews and meta-analyses: the PRISMA statement. Annals of internal medicine. 2009 Aug 18;151(4):264–9.

8. Open Meta Analyst: Wallace, Byron C., Issa J. Dahabreh, Thomas A. Trikalinos, Joseph Lau, Paul Trow, and Christopher H. Schmid. “Closing the Gap between Methodologists and End-Users: R as a Computational Back-End.” Journal of Statistical Software 49 (2012): 5.”

9. Higgins JP, Thompson SG, Deeks JJ, Altman DG. Measuring inconsistency in meta-analyses. Bmj. 2003 Sep 4;327(7414):557–60.

10. Wallace BC, Lajeunesse MJ, Dietz G, Dahabreh IJ, Trikalinos TA, Schmid CH, Gurevitch J. Open MEE: Intuitive, open‐source software for meta‐analysis in ecology and evolutionary biology. Methods in Ecology and Evolution. 2017 Aug;8(8):941–7.

11. Stangl D, Berry DA, editors. Meta-analysis in medicine and health policy. CRC Press; 2000 Apr 20.

12. Egger M, Davey-Smith G, Altman D, editors. Systematic reviews in health care: meta-analysis in context. John Wiley & Sons; 2008 Apr 15.

13. Hirsch IB, Bode BW, Garg S, Lane WS, Sussman A, Hu P, Santiago OM, Kolaczynski JW. Continuous subcutaneous insulin infusion (CSII) of insulin aspart versus multiple daily injection of insulin aspart/insulin glargine in type 1 diabetic patients previously treated with CSII. Diabetes Care. 2005 Mar 1;28(3):533–8.

14. Bolli GB, Kerr D, Thomas R, Torlone E, Sola-Gazagnes A, Vitacolonna E, Selam JL, Home PD. Comparison of a multiple daily insulin injection regimen (basal once-daily glargine plus mealtime lispro) and continuous subcutaneous insulin infusion (lispro) in type 1 diabetes: a randomized open parallel multicenter study. Diabetes care. 2009 Jul 1;32(7):1170–6.

15. Hanaire-Broutin HÉ, Melki V, Bessières-Lacombe SY, Tauber JP. Comparison of continuous subcutaneous insulin infusion and multiple daily injection regimens using insulin lispro in type 1 diabetic patients on intensified treatment: a randomized study. The Study Group for the Development of Pump Therapy in Diabetes. Diabetes Care. 2000 Sep 1;23(9):1232–5.

16. Alemzadeh R, Ellis JN, Holzum MK, Parton EA, Wyatt DT. Beneficial effects of continuous subcutaneous insulin infusion and flexible multiple daily insulin regimen using insulin glargine in type 1 diabetes. Pediatrics. 2004 Jul 1;114(1):e91–5.

17. Skogsberg L, Fors H, Hanas R, Chaplin JE, Lindman E, Skogsberg J. Improved treatment satisfaction but no difference in metabolic control when using continuous subcutaneous insulin infusion vs. multiple daily injections in children at onset of type 1 diabetes mellitus. Pediatric diabetes. 2008 Oct;9(5):472–9.

18. Weintrob N, Schechter A, Benzaquen H, Shalitin S, Lilos P, Galatzer A, Phillip M. Glycemic patterns detected by continuous subcutaneous glucose sensing in children and adolescents with type 1 diabetes mellitus treated by multiple daily injections vs continuous subcutaneous insulin infusion. Archives of pediatrics & adolescent medicine. 2004 Jul 1;158(7):677–84.

19. Reznik Y, Cohen O, Aronson R, et al.; OpT2mise Study Group. Insulin pump treatment compared with multiple daily injections for treatment of type 2 diabetes (OpT2mise): a randomized open label controlled trial. Lancet 2014;384:1265–1272

20. Doyle EA, Weinzimer SA, Steffen AT, Ahern JA, Vincent M, Tamborlane WV. A randomized, prospective trial comparing the efficacy of continuous subcutaneous insulin infusion with multiple daily injections using insulin glargine. Diabetes Care. 2004;27(7):1554–8.

21. Lepore G, Dodesini AR, Nosari I, Trevisan R. Effect of continuous subcutaneous insulin infusion vs multiple daily insulin injection with glargine as basal insulin: an open parallel long-term study. Diabetes, nutrition & metabolism. 2004 Apr;17(2):84–9.

22. Marshall SM, Home PD, Taylor R, Alberti KGMM. Continuous subcutaneous insulin infusion versus injection therapy: a randomized cross-over trial under usual diabetic clinic conditions. Diabetic Med 1987;4:521–5.

23. American Diabetes Association. 7. Diabetes Technology: Standards of Medical Care in Diabetes—2020. Diabetes Care. 2020 Jan 1;43(Supplement 1):S77–88.

24. Aziz KMA. Management of type-1 and type-2 diabetes by insulin injections in diabetology clinics – A scientific research review. Recent patents on endocrine, metabolic & immune drug discovery. 2012 May 1;6(2):148–70.

